# Effects of an 18-month meditation training on subjective and objective sleep quality in older adults: a secondary analysis of the Age-Well randomized controlled trial

**DOI:** 10.64898/2026.09.30.26364409

**Authors:** Pierre Champetier, Anaïs Hamel, Stéphane Rehel, Françoise Bertran, Oriane Hebert, Marion Delarue, Gaël Chetelat, Antoine Lutz, Alison Mary, Géraldine Rauchs, the Medit-Ageing Research Group

## Abstract

**Study objectives:** Aging is associated with increased sleep complaints and changes in sleep architecture, microstructure and rest-activity rhythms. Meditation-based interventions have shown benefits on subjective sleep quality, but their effects on objective sleep parameters remain largely underexplored. This study investigated the effects of an 18-month meditation intervention on sleep combining subjective (questionnaires) and objective (polysomnography and actigraphy) measures in older adults.

**Methods:** A total of 120 cognitively unimpaired older adults (≥65 y) with minimal sleep complaint completed questionnaires, one week of actigraphy and in-home polysomnography at baseline, and again 18 months later. Following the baseline visit, participants were randomly assigned to one of 3 groups: meditation training, non-native language training (active control), or passive control. Group-by-visit interactions were examined using mixed-effects models for sleep questionnaires, actigraphy-derived metrics, sleep architecture and microstructure, including slow waves, spindles, and spectral power.

**Results:** Compared to the passive control group only, meditation training was associated with greater ease of morning awakening (p=0.04). No significant intervention effects were observed for any other sleep parameter, including actigraphy- and polysomnography-derived measures. These findings remained unchanged after adjustment for depressive symptoms and trait anxiety.

**Conclusion:** In cognitively unimpaired older adults, this 18-month meditation intervention yielded only limited improvements in subjective sleep assessment. Meditation may therefore have subtle effects on self-reported sleep quality in this healthy older population, although no effects on objective sleep measures were detected. Future studies should examine whether adapting the content, intensity, or timing of meditation practice and enrolling populations with clinically meaningful sleep disturbances or psychoaffective symptoms may yield greater benefits.

**Statement of significance:** Sleep quality is impaired with age and may contribute to cognitive decline. This randomized controlled trial is the first to assess the effects of an 18-month meditation intervention on both subjective and objective sleep parameters in cognitively unimpaired older adults, compared with active and passive control groups. Meditation improved perceived ease of morning awakening compared to the passive control group, but no other group differences were observed. These findings suggest that meditation, an accessible and low-cost intervention, may modestly improve some aspects of self-reported sleep quality, while having limited effects on objective sleep measures in older adults without sleep complaints. Studies evaluating the effects of meditation on objective sleep measures in more diverse and clinically affected populations are needed.

**Graphical abstract:** 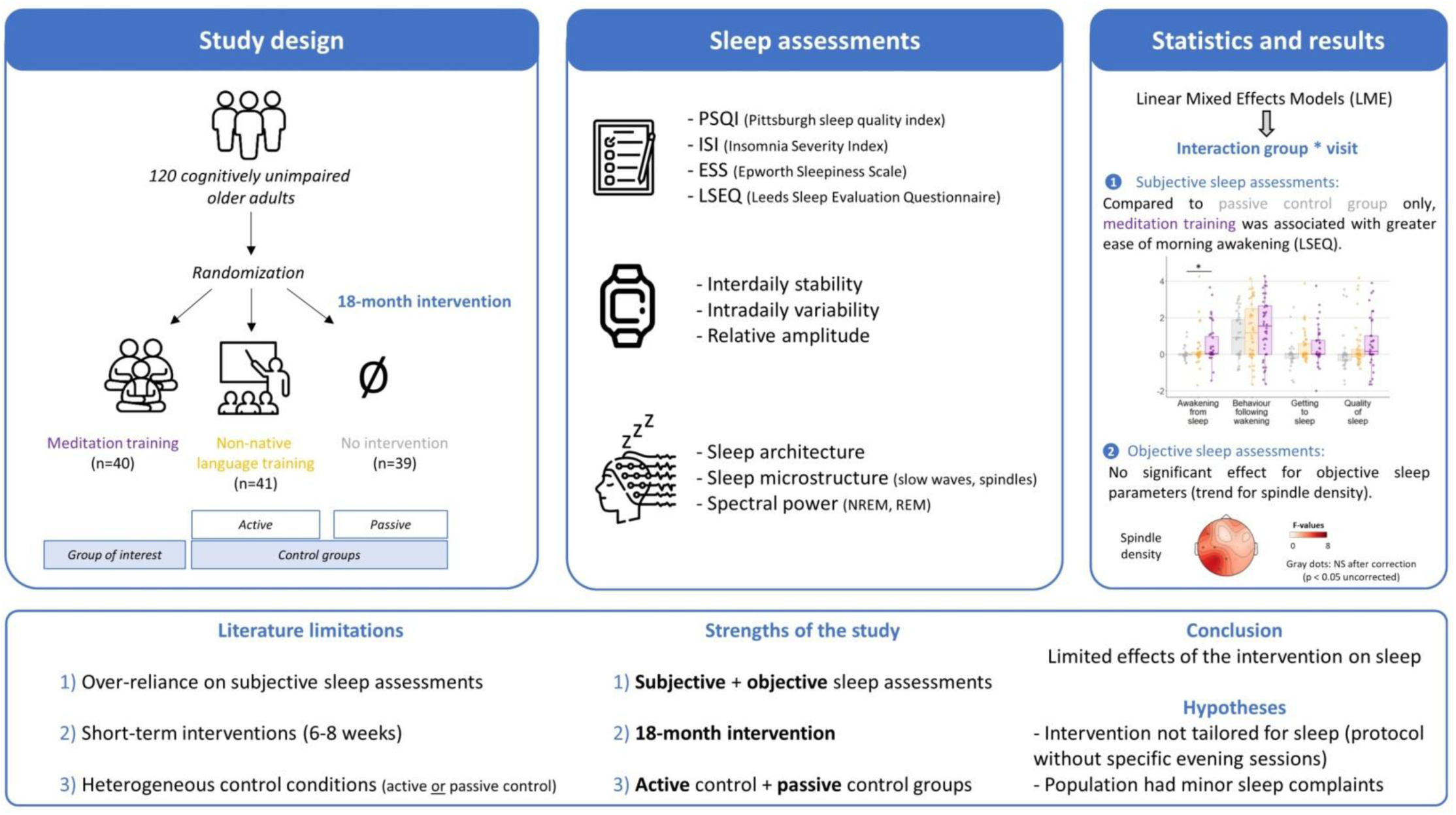

## Introduction

Aging is associated with alterations in sleep quality, which can significantly impact global health and well-being. Around 50% of individuals aged over 65 experience difficulties initiating or maintaining sleep ^1,2^. Older adults also show changes in rest-activity rhythms, including earlier sleep timing (i.e., earlier bedtime and rise time), reduced circadian amplitude, and greater daily fragmentation ^3,4^. Moreover, advancing age is associated with longer sleep latency, more fragmented sleep, a greater proportion of lighter sleep stage (N1), and less deep sleep (slow wave sleep or N3), resulting in less restorative sleep ^4,5^. These sleep disturbances are increasingly recognized as potential risk factors for cognitive decline and dementia, including Alzheimer’s disease ^6^. Some studies further suggest that sleep disturbances occurring during midlife may predict subsequent cognitive decline and contribute to neurodegeneration ^7^. Developing non-pharmacological interventions aimed at improving sleep may be a relevant strategy for promoting healthy aging.

In this context, meditation-based training is a particularly promising approach ^8,9^. Indeed, meditation training has been linked to cognitive benefits, with studies showing improvements in attentional and emotional self-regulation, as well as structural and functional changes in brain regions involved in these processes, including frontal and limbic areas, as well as the insula. It has also been associated with reduced psychoaffective symptoms (such as depression, stress and anxiety) that are known to exacerbate age-related sleep alterations ^8–10^. Meditation training may influence sleep through multiple pathways. Notably, it may reduce anxiety and depression, which may give rise to maladaptive mechanisms like ruminations that contribute to sleep disturbances. It may also enhance cognitive and emotional regulation mechanisms that support overall well-being. Two recent meta-analyses indicate that 6 or 8 weeks of mindfulness-based meditation training can improve subjective sleep quality in older adults with sleep disturbances ^11,12^. Most studies in this field have focused on older individuals with clinically-relevant sleep complaints. In these populations, mindfulness-based training improved subjective sleep quality, particularly insomnia symptoms, compared to a passive wait-list control group ^13^, or a sleep hygiene education program ^14,15^. However, no significant differences have been reported in subjective sleep quality assessed using the Pittsburgh Sleep Quality Index (PSQI) when mindfulness-based training was compared with other active interventions (e.g., cognitive fitness training or combined sleep hygiene, education, and physical activity programs) ^15,16^.

Studies including objective sleep measures are scarcer. One randomized controlled trial (RCT) compared the effect of mindfulness-based training to a program combining sleep hygiene, education, and physical activity on polysomnography-derived metrics (sleep architecture and electroencephalography (EEG) spectral power). No significant interaction between group and time favoring mindfulness training was observed in older adults with sleep complaints ^17^.

Overall, the literature examining the effects of meditation interventions on sleep in older adults has several limitations. First, most studies rely exclusively on self-report questionnaires, leaving objective sleep measures largely unexplored. Consequently, key sleep parameters that are particularly affected by aging, such as slow waves and sleep spindles, have not yet been investigated in the context of meditation interventions. Second, interventions are typically limited to 6- or 8-week training programs, which may be insufficient to produce measurable changes in sleep. Third, the heterogeneity of control conditions, whether active or passive, limits the ability to isolate meditation-specific effects on sleep quality. As a result, it remains unclear whether the observed improvements reflect the specific mechanisms of mindfulness practice or non-specific effects associated with participation in an intervention. Fourth, previous studies have primarily targeted older adults with self-reported sleep disturbances, with relatively little attention given to those with minimal sleep complaints. Finally, previous studies have focused solely on mindfulness meditation, whereas other practices, such as loving-kindness and compassion meditation, remain largely unexplored. These practices may complement mindfulness by enhancing positive emotions, fostering a greater sense of purpose in life, and strengthening social support ^18,19^. Integrating mindfulness, loving-kindness and compassion meditation within a single intervention may therefore enhance the benefits of meditation for healthy aging ^9^.

Addressing these methodological limitations is essential for accurately determining the effectiveness of meditation as an approach to improve sleep in older adults. The present study, part of the Age-Well RCT ^20,21^, investigated the impact of an 18-month meditation intervention, the longest ever tested to date in older adults, on sleep in cognitively healthy older individuals. Sleep was evaluated using complementary subjective measures (questionnaires) and objective assessments (actigraphy and polysomnography). We compared meditation training to a matched non-native language training program (active control) and a no-intervention condition (passive control). The assessment encompasses self-reported sleep quality across several dimensions, rest-activity rhythm, sleep architecture, and sleep microstructure (slow waves, sleep spindles, and EEG spectral power). We hypothesized that meditation training would lead to greater improvements in both subjective and objective sleep quality than both control conditions, particularly for sleep parameters commonly altered with aging, including insomnia symptoms, sleep latency, sleep duration and efficiency, and slow-wave sleep.

## Method

### Participants and study design

Participants were included in the Age-Well randomized clinical trial (RCT) ^21^, sponsored by the French National Institute of Health and Medical Research (INSERM). The general design of the trial including details on randomization, masking of treatment allocation, interventions and sample size calculation have been described in detail previously ^21^. Briefly, 137 cognitively unimpaired older adults were recruited from the general population and met the following main eligibility criteria: native French speaker, 65 years old or older, retired for at least one year, with a minimum of 7 years of formal education and showing performance within the normal range for their age and educational level on standardized cognitive tests (for detailed criteria, refer to Poisnel et al., 2018 ^21^). Exclusion criteria included any major neurological or psychiatric disorders, chronic or acute unstable illnesses, a history of cerebrovascular disease, and the use of medications that may interfere with cognitive functioning. Following the baseline visit, participants were randomized (1:1:1) to (i) an 18-month meditation training intervention group, (ii) an 18-month non-native language (English) training intervention group (active control), or (iii) a no-intervention group (passive control). Pre- and post-intervention visits comprised multimodal assessments including, among other biological and neuroimaging examinations not detailed here, a comprehensive neuropsychological assessment, complemented by questionnaires to assess depressive symptoms (Geriatric Depression Scale, GDS ^22^), trait anxiety (State-Trait Anxiety Inventory B, STAI-B ^23^), and sleep, as well as, actigraphy and polysomnography. All data were collected within a maximum period of 3 months. **Figure 1** shows a flow diagram of the inclusion process. Of the 137 randomized participants, 120 were analyzed in the present study (mean age ± SD: 69.2 ± 3.7 years, 63% women). Two participants were excluded due to major eligibility criteria not met (i.e., history of head trauma and multiple sclerosis diagnosis after an intermediate 9-month visit [with a likely subclinical state at inclusion]), 1 died during the follow-up and 14 participants were treated by continuous positive airway pressure (CPAP) due to obstructive sleep apnea at baseline, during the intervention or at post-intervention. One participant revealed not having followed his allocated arm (no intervention), and instead attended the non-native language training. Consequently, the participant was analyzed within the non-native language training group. All participants provided written informed consent before undergoing any examination. The Age-Well randomized clinical trial was approved by the ethics committee (Comité de Protection des Personnes Nord-Ouest III, Caen, France; trial registration number: EudraCT: 2016-002441-36; IDRCB: 2016-A01767-44; ClinicalTrials.gov Identifier: NCT02977819).

**Figure 1.**
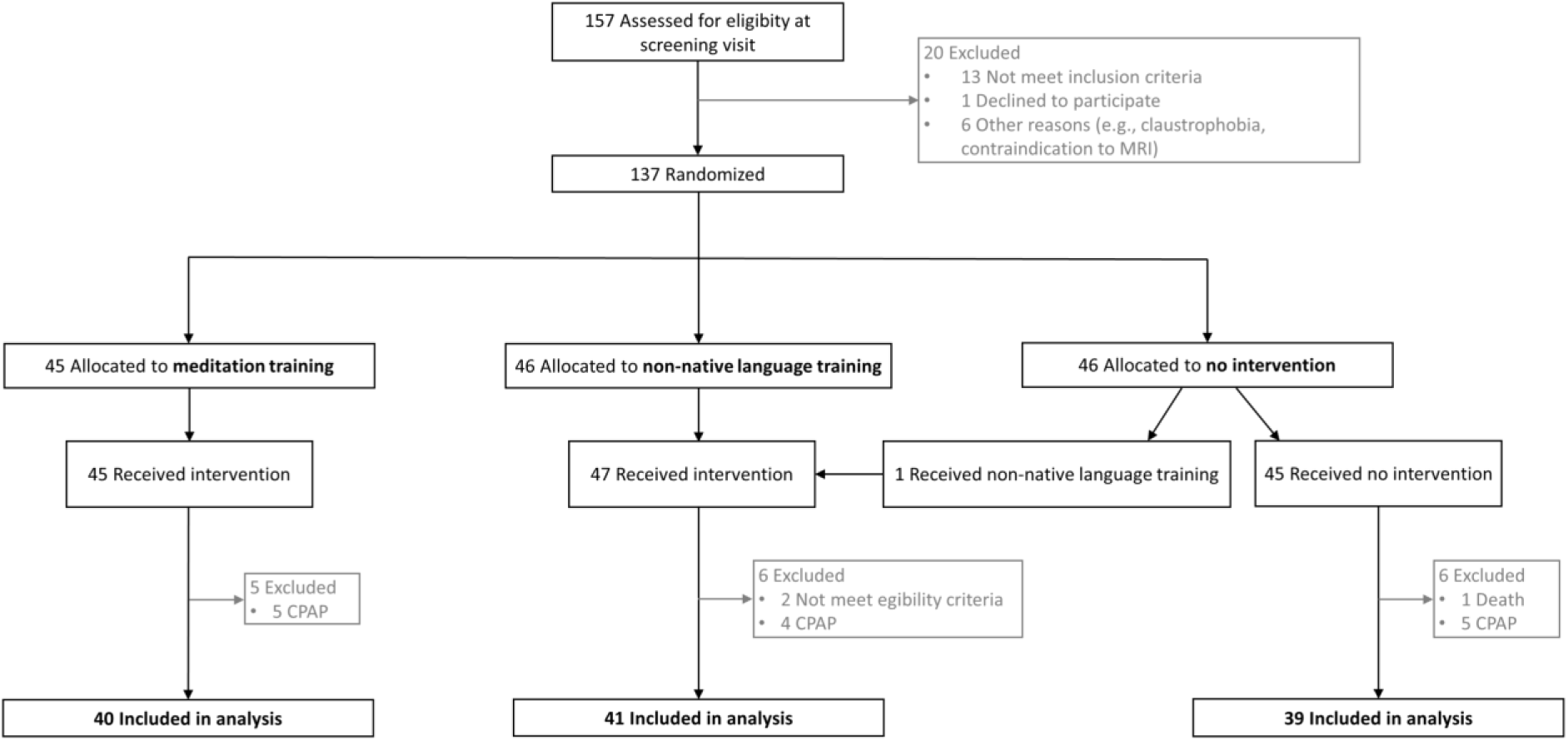
CONSORT diagram of the Age-Well study enrolment and randomization.

### Interventions

The interventions have been fully described previously ^20,21^. The meditation and non-native language training interventions were structurally equivalent in overall course length, class time, and home activities. Both programs lasted 18 months and included weekly 2-hour group sessions, daily home practice (at least 20 minutes), and one day of intensive practice (5 hours) during the intervention period.

The meditation training consisted of an original secular program of meditation training called “The Silver Sante Study Meditation Program”, specially developed for this study based on pre-existing interventions for personal development and healthy aging. It was delivered in Caen (France) by experienced meditation instructors. The aim of this 18-month intervention program was to develop mindfulness, kindness and compassion abilities as psychological resources to help participants cope with the physical, cognitive and psychosocial challenges of aging ^9^. It comprised 9 months of mindfulness meditation practice, followed by 9 months of loving kindness and compassion meditation practice. Sessions included seated or walking meditation, teaching components, and group discussions.

The non-native language training program was designed as a cognitively stimulating active control intervention for native French speakers. It consisted of English classes aimed at strengthening participants’ abilities to understand, write, and speak English. Sessions were tailored to participants’ baseline English proficiency and included activities designed to promote the acquisition of vocabulary and grammatical structures.

Participants in the no-intervention group were asked to maintain their habits throughout the 18-month study period. They were specifically asked not to engage in meditation or non-native language training during this period.

### Sleep assessment

#### Questionnaires

Three questionnaires were used to assess self-reported sleep and daytime sleepiness: the PSQI ^24^, the Insomnia Severity Index (ISI) ^25^, and the Epworth Sleepiness Scale (ESS) ^26^. A PSQI total score > 5 was considered indicative of poor sleep quality and suggestive of sleep complaints. Moreover, intervention-related changes in sleep and wakefulness were evaluated using the Leeds Sleep Evaluation Questionnaire (LSEQ) ^27^. The LSEQ comprises ten 100-mm self-rating visual analogue scales to assess sleep quality, and has demonstrated good test-retest reliability ^27,28^. The LSEQ assesses four dimensions of sleep and morning functioning: ease of getting to sleep (GTS, items 1,2,3), quality of sleep (QOS, items 4,5), awakening from sleep (AFS, items 6,7), and behavior following wakefulness (BFW, items 8,9,10). Mean scores were calculated for each dimension.

#### Actigraphy

The rest-activity cycle was recorded using the MotionWatch 8 wrist-worn triaxial actigraph (CamNTech Ltd, Cambridge, UK), for at least six consecutive nights (mean = 7.66 ± 0.51; range = [6:8] nights). Participants were instructed to continuously wear the device on their nondominant wrist until the end of the recording. They had to press the event marker button at lights off and lights on, and to fill in a sleep diary each morning to facilitate data analysis.

Data were analyzed using the MotionWare software (version 1.1.25, CamNTech Ltd, Cambridge, UK) and sampled using a 5-second epoch. A sensitivity threshold of 20 counts was applied to distinguish activity from rest. For each night, the “Lights Off” and “Got Up” markers were placed in the appropriate position following the event marker put by the participant and cross validated with the light sensor data and sleep diary information. Then, “Fell Asleep” and “Woke up” markers were automatically adjusted from the combination of the activity data and the “Lights Off” and “Got Up” markers, respectively, allowing the collection of the assumed sleep time. Rest-activity rhythm variables included interdaily stability, intradaily variability, which quantify the degree of regularity in the activity-rest patterns across days and the degree of fragmentation of activity-rest periods respectively ^29^. Relative amplitude, which measures the relative difference between the mean activity during the 10 most active hours and the 5 least active ones, was also assessed.

#### Polysomnography

In-home polysomnography was recorded using a portable recording device (Siesta®, Compumedics, Australia). Twenty EEG electrodes were positioned over the scalp according to the international 10-20 system (Fp1, Fp2, F3, F4, F7, F8, FZ, C3, C4, CZ, T3, T4, P3, P4, PZ, O1, O2, vertex ground and bi-mastoid reference), with impedances kept below 5 kΩ. The EEG signal was digitalized at 256 Hz. Respiratory movements, airflow and oxygen saturation were also monitored to detect respiratory events during sleep. Polysomnography recordings were visually scored in 30-second epochs following the American Academy of Sleep Medicine’s international scoring guidelines ^30^. Conventional polysomnography parameters of sleep were obtained, which include sleep latency, total sleep time (TST), wake after sleep onset (WASO), sleep efficiency (100 x time asleep/TST), the proportion of each sleep stage (N1, N2, N3 and REM sleep, expressed as % of TST) and the apnea-hypopnea index (AHI, number of respiratory events/hour of sleep).

#### EEG preprocessing

EEG preprocessing, detection of sleep oscillations and power analyses were conducted using Matlab and the Sleeptrip toolbox (https://github.com/Frederik-D-Weber/sleeptrip; RRID: SCR_017318) based on FieldTrip functions (http://fieldtriptoolbox.org; RRID: SCR_004849). High-pass and low-pass filters were applied at 0.3 Hz and 50 Hz, respectively. In line with recent recommendations for sleep research in older adults, we considered N2 and N3 sleep stages as a continuum and performed the analyses on these combined stages. Analyses of NREM sleep (i.e., N2 and N3) and REM sleep were performed separately. For all subjects, an independent component analysis (ICA) was performed on REM epochs to remove ocular (and more occasionally ECG artefacts) on EEG signals. The EEG data of all participants were visually checked after ICA. Data were then epoched into 5-s bins. All recordings were visually inspected and the 5-s epochs containing EEG artefacts or arousals were rejected. Channels with artefacts affecting a majority of the recording were removed. If all channels within a specific region were removed due to artefacts, data from that region were unavailable for the participant concerned. Consequently, the number of participants included for spectral analyses varies between regions (see Supplementary Material).

#### Slow wave and spindle detection

Slow waves and sleep spindles were automatically detected in artifact-free NREM sleep epochs using a validated algorithm adapted from a previous study ^31^. Slow wave detection was carried out after applying high-pass and low-pass filters with cutoff frequencies of 0.5 Hz and 3.5 Hz, respectively (−3 dB cutoff, two-pass 4th order IIR Butterworth). Consecutive positive-to-negative zero crossings were identified, and those with durations corresponding to a frequency between 0.5 and 1.11 Hz were marked as potential slow waves. Putative slow waves were discarded if they had a negative down-peak greater than -15 µV, a positive up-peak less than 10 µV, or an amplitude greater than 600 µV. Slow waves were kept based on individually adjusted threshold: a down-peak potential less than 1.25 of the mean down-peak potential and an amplitude greater than 1.25 of the mean amplitude of all other putative slow waves within this channel ^32^. For each participant and channel, the mean amplitude, duration, and density (number of slow waves per artifact-free minute of N2 and N3 sleep) were calculated.

To detect spindles, the EEG signal was band-pass filtered between 11 and 15 Hz. The root mean square (RMS) signal was computed across the entire recording, and then smoothed with a moving average using a 0.2-second window. Spindles were identified when the smoothed RMS signal exceeded an individual amplitude threshold (1.5 times the standard deviation of the filtered signal) for a duration between 0.5 and 2 seconds. Spindles with amplitudes greater than 120 μV were excluded. For each participant and channel, the mean peak-to-trough amplitude, duration, and spindle density (number of spindles per artifact-free minute of N2 and N3 sleep) were calculated for each electrode.

#### Spectral power analyses

EEG spectral power was calculated for NREM sleep and REM sleep separately. Power spectral analyses were performed on continuous 4-second segments with a 50% overlap and were tapered using a Hanning window. A fast Fourier Transform was then applied. The power spectra were averaged using Welch’s method and normalized based on the effective noise bandwidth. For each electrode, mean power was computed across specific frequency bands: delta (0.5-4 Hz), theta (4-8 Hz), alpha (8-12 Hz), sigma (12-15 Hz) and beta (15-25 Hz) frequency bands during NREM. Absolute EEG spectral power values were normalized by the total spectral power for each epoch, corresponding to the relative spectral power.

### Statistical analysis

Differences in demographic characteristics between groups at baseline were assessed using ANOVA, followed by Tukey post hoc tests for continuous variables, and χ² tests for categorical variables. Intervention-related subjective sleep changes between groups, as assessed using the LSEQ, were analyzed using ANCOVA when statistical assumptions were met. If these assumptions were not satisfied, Kruskal–Wallis H tests were applied. Post hoc pairwise comparisons were performed only when the main effect of group was statistically significant, using Dunn post hoc tests with Bonferroni correction for multiple comparisons. In the LSEQ analysis, one participant was excluded due to missing data. Statistical analyses of the remaining questionnaires and sleep architecture were conducted using a modified intention-to-treat approach, in which participants were analyzed based on their assigned group, except the participant who deviated from the assigned intervention and was analyzed according to the treatment received. Depending on the distribution of the dependent variable, linear and generalized linear mixed-effects models with random intercepts at the participant level were used to analyze each sleep parameter. The effects of the intervention were assessed by examining the interaction between trial arm and visit (Group × Visit). Log transformation was applied if necessary to normalize the data and achieve a normal distribution. Mixed-effects models were controlled for age, sex, education level, AHI, STAI-B score (anxiety trait), and GDS score (depressive symptoms), with continuous variables being mean-centered. Residuals from the statistical models were inspected for potential outliers (≥ ±3 standard deviations), any identified outliers were removed. When the Group × Visit interaction was statistically significant, post hoc pairwise comparisons were performed, adjusting for multiple comparisons (using the Tukey method). Analyses were performed using R version 4.1.1 (R Foundation for Statistical Computing). The *lme4* package was used for mixed-effects models, the *ANOVA* function for omnibus tests, and the *emmeans* package for pairwise comparisons and estimated marginal means. All statistical models satisfied the corresponding assumptions. The significance level adjusted for multiple comparisons was set at p <0.05. Statistical analyses of all EEG-derived metrics computed at each electrode (i.e., slow waves, spindles and EEG power) were performed using linear mixed-effects models combined with a cluster-based permutation approach derived from Maris & Oostenveld ^33^ with Matlab R2018b. For each electrode, the F- and p-values for the Group × Visit interaction were calculated. Candidate clusters were defined as groups of at least two neighboring electrodes with p < 0.025 (cluster alpha). For each candidate cluster, the cluster statistic was computed as the sum of the F-values across all electrodes within the cluster. To generate the null distribution, group labels were randomly permuted 1,000 times. For each permuted dataset, the candidate cluster with the largest absolute cluster statistic was retained. The cluster statistics obtained from the original dataset were then compared with this null distribution to derive Monte Carlo p-values. Clusters with p_cluster_ < 0.05 were considered statistically significant, indicating that the observed cluster statistic fell within the most extreme 5% of the permutation distribution (i.e., below the 5th percentile for negative clusters or above the 95th percentile for positive clusters).

## Results

### Baseline characteristics

Demographic characteristics and psychoaffective factors at baseline are summarized for each group in supplementary **Table S1**. Despite a marginal effect of group on age (F(2,117) = 3.097, p = 0.049), post hoc pairwise comparisons did not reveal any significant age difference between the three groups. Education level was statistically different between groups (F(2,117) = 3.51, p = 0.03). Post hoc tests revealed that the no-intervention group was more educated than the non-native language training group ([95% CI]: 1.76 [0.18, 3.34], p = 0.02, Tukey-corrected).

### Sleep questionnaires

Scores on sleep questionnaires for each group and visit are reported in supplementary **Table S2**. Approximately 37% of the subjects had a PSQI score higher than 5, the threshold commonly used to distinguish individuals with and without sleep disturbances. The analysis of the LSEQ, a questionnaire specifically designed to assess perceived changes in sleep following an intervention, indicated a significant between-group difference in the change in the “Awakening from Sleep” dimension relative to baseline (H(2) = 6.53, p = 0.04; **Table 1**). Specifically, post hoc analyses revealed that the meditation training group showed greater improvement in this dimension compared to the no-intervention group (z = 2.49, p = 0.04, Bonferroni-corrected), whereas the other pairwise comparisons were not significant (**Table 1**, **Figure 2**). Analysis of the Group × Visit interaction for the other sleep questionnaires (PSQI, ESS, and ISI) revealed no additional statistically significant effects (**Table 2**).

**Table 1.** Group differences in the Leeds Sleep Evaluation Questionnaire (LSEQ).

|  | Omnibus test | Meditation training<br>vs.<br>Non-native language training |  | Meditation training<br>vs.<br>No-intervention |  | Non-native language training<br>vs.<br>No-intervention |  |
| --- | --- | --- | --- | --- | --- | --- | --- |
| LSEQ | Test statistic,<br>p-value | Test Statistic | p-value | Test Statistic | p-value | Test Statistic | p-value |
| Awakening<br>from sleep | **H(2) = 6.53,**<br>**p = 0.04** | z = -1.72 | p<sub>adj</sub> = 0.26 | **z = -2.49** | **p<sub>adj</sub> = 0.04** | z = -0.81 | p<sub>adj</sub> = 1 |
| Behavior<br>following<br>wakefulness | F(2,105) = 1.02,<br>p = 0.38 | NA | NA | NA | NA | NA | NA |
| Getting to sleep | H(2) = 5.25,<br>p = 0.07 | NA | NA | NA | NA | NA | NA |
| Quality of sleep | H(2) = 4.13,<br>p = 0.13 | NA | NA | NA | NA | NA | NA |

*Note: ANCOVA and Kruskal-Wallis models were used to assess group differences in the Leeds Sleep Evaluation Questionnaire (LSEQ). Post hoc pairwise comparisons were performed only when the main effect of group was statistically significant. Dunn's test was used following significant Kruskal–Wallis tests. Adjusted p-values (p<sub>adj</sub>) are reported. Abbreviations: LSEQ (Leeds Sleep Evaluation Questionnaire), NA (Not Applicable).*

**Figure 2.**
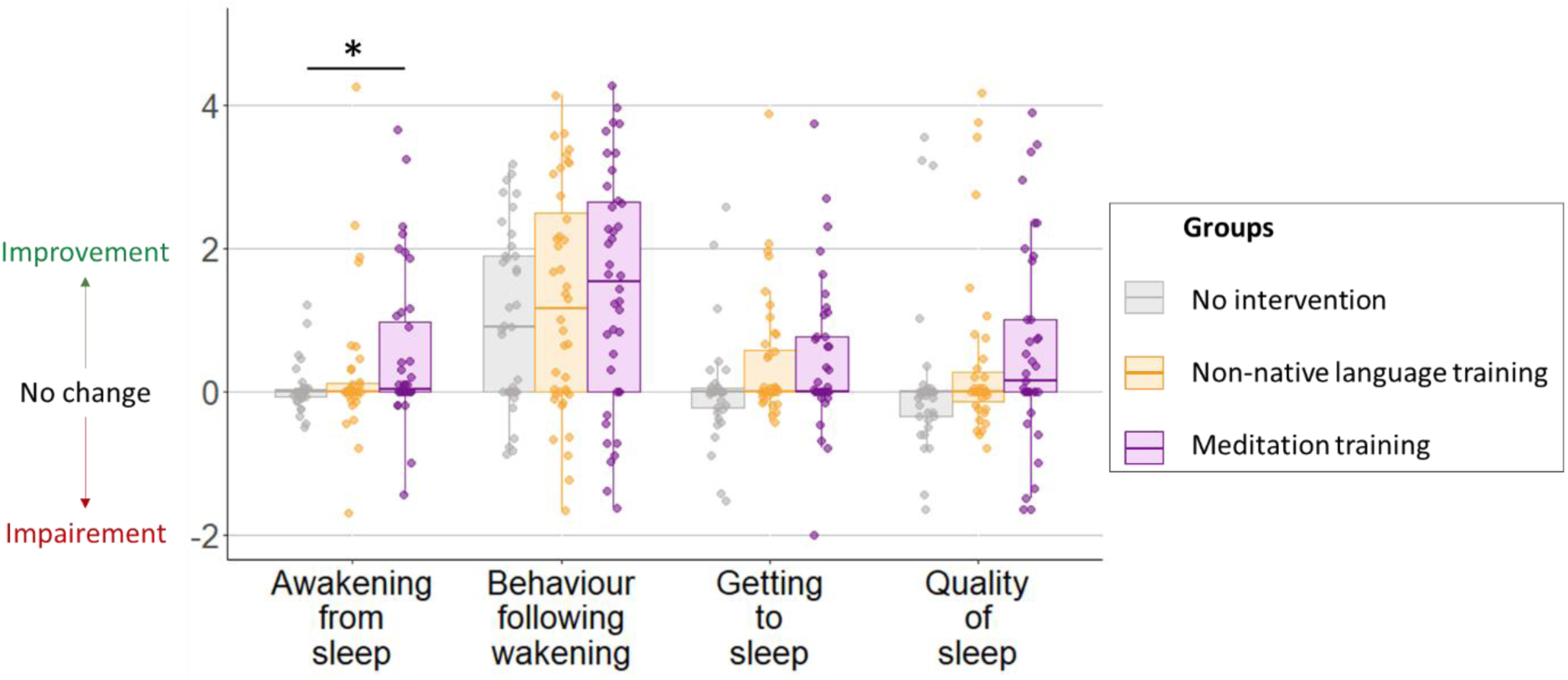
Subjective assessment of changes in sleep quality using the Leeds Sleep Evaluation Questionnaire (LSEQ) after 18 months of intervention in each group. Analyses revealed an improvement of “awakening from sleep” for the meditation training group compared to the no-intervention group. * p < 0.05. This item was assessed through a self-administered questionnaire asking participants whether this aspect had improved or deteriorated compared to before the intervention, using ten 100-mm visual analogue scales.

**Table 2.** Group × Visit interaction effects for sleep questionnaire scores, actigraphy-derived metrics, and standard polysomnography-derived metrics.

| Interaction Group × Visit | F-value<br>or<br>$\chi^2$-value | p-value |
| --- | --- | --- |
| **Sleep questionnaires** |  |  |
| PSQI | F(2,107.1) = 0.27 | 0.76 |
| ISI | F(2,101.9) = 0.30 | 0.74 |
| ESS | F(2,107.3) = 0.23 | 0.79 |
| **Rest-activity rhythm** |  |  |
| Interdaily stability | F(2,87.7) = 0.85 | 0.43 |
| Intradaily variability | F(2,103.9) = 0.78 | 0.46 |
| Relative amplitude | $\chi^2(2) = 0.51$ | 0.77 |
| **Sleep architecture** |  |  |
| TST (min) | F(2,113.6) = 2.94 | 0.06 |
| Sleep efficiency (%) | $\chi^2(2) = 5.87$ | 0.05 |
| Sleep latency (min) | F(2,116.2) = 2.69 | 0.07 |
| WASO (min) | F(2,112.8) = 1.32 | 0.27 |
| NREM-1 (% TST) | F(2,113.3) = 0.02 | 0.98 |
| NREM-2 (% TST) | F(2,108.6) = 0.01 | 0.99 |
| NREM-3 (% TST) | F(2,110.6) = 0.19 | 0.83 |
| REM (% TST) | F(2,112.2) = 1.02 | 0.36 |

*Linear mixed-effects models (LMMs) are reported with F statistics, while generalized linear mixed-effects models (GLMMs) are reported with $\chi^2$ statistics. All analyses were corrected for age, sex, education level, AHI, STAI-B score, and GDS score. The Group × Visit interaction tests whether changes over time differ between the three groups. Abbreviation: CI (Confidence Interval), NREM (Non-rapid eye movement sleep), REM (rapid eye movement sleep), TST (Total Sleep Time), WASO (wake after sleep onset).*

### Rest-activity rhythms and sleep architecture

Rest-activity rhythm and sleep architecture parameters for each group and visit are reported in supplementary **Tables S3 and S4**. Changes in rest-activity rhythms parameters (interdaily stability, intradaily variability and relative amplitude) over time did not differ significantly between the three groups (all p > 0.05, **Table 2**). Similarly, the Group × Visit interaction was not significant on conventional parameters of sleep architecture, namely sleep latency, total sleep time (TST), wake after sleep onset (WASO), sleep efficiency, and the proportion of each sleep stage (all p ≥ 0.05, **Table 2**).

### Sleep microstructure and spectral power

The Group × Visit interaction was tested at each channel for NREM sleep oscillations (slow waves and spindles) and for EEG power during NREM and REM sleep. Although p-values below 0.05 were observed at the channel level for a few EEG channels (notably for spindle density), no analysis yielded a significant cluster following the cluster-based permutation approach (**Figures 3 and 4**). Between-group changes in spindle density over centroparietal derivations, after 18 months of intervention in each group are shown in Supplementary **Figure S1**.

**Figure 3.**
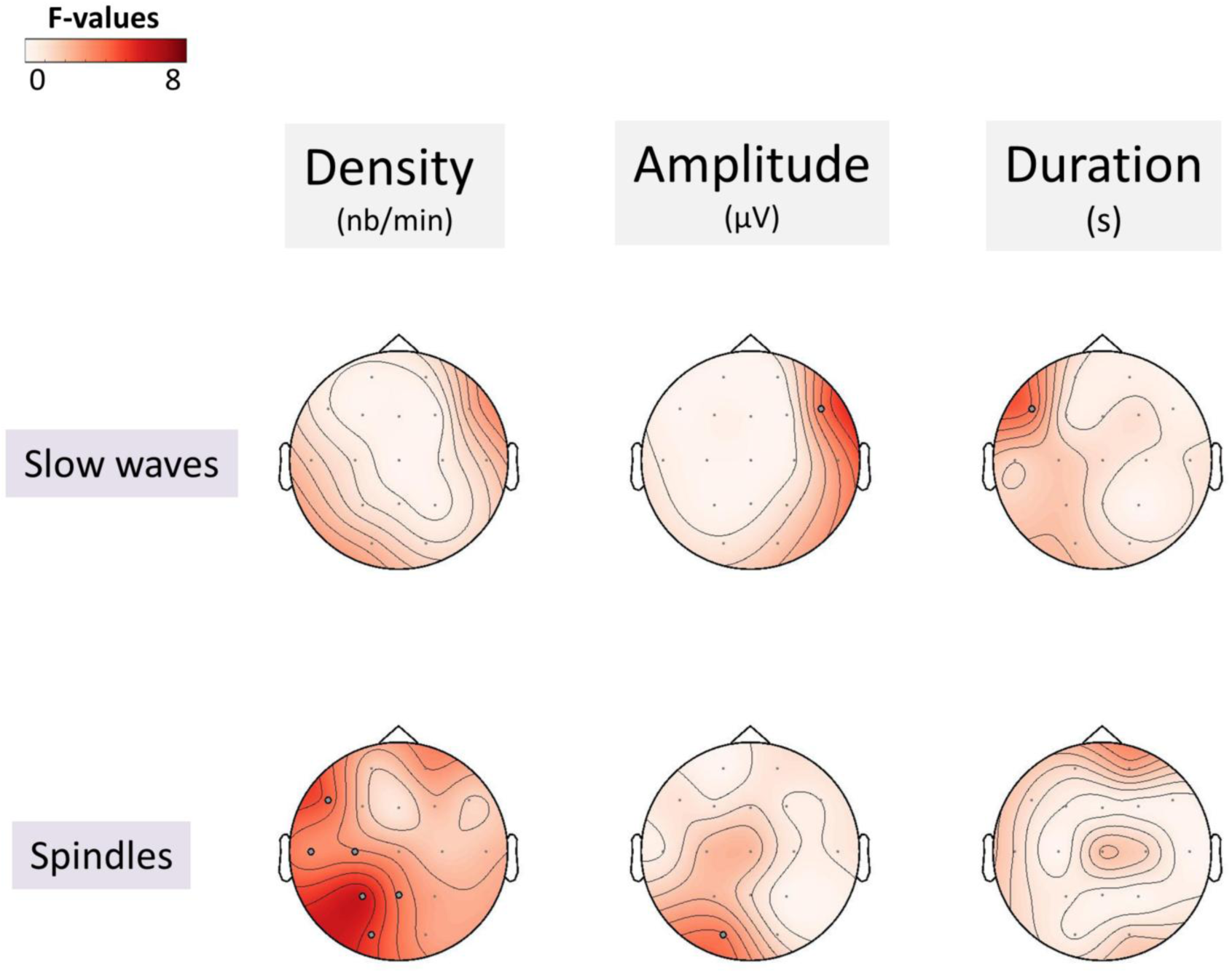
Between-group differences in changes in NREM sleep oscillations following the intervention. Statistical maps (F-values) of the Group × Visit interaction effects on each parameter (density, amplitude, and duration) of NREM sleep oscillations (slow waves and spindles). Gray dots (•) indicate channels with p-value < 0.05 at the channel level that did not reach significance after applying the cluster-based permutation approach.

**Figure 4.**
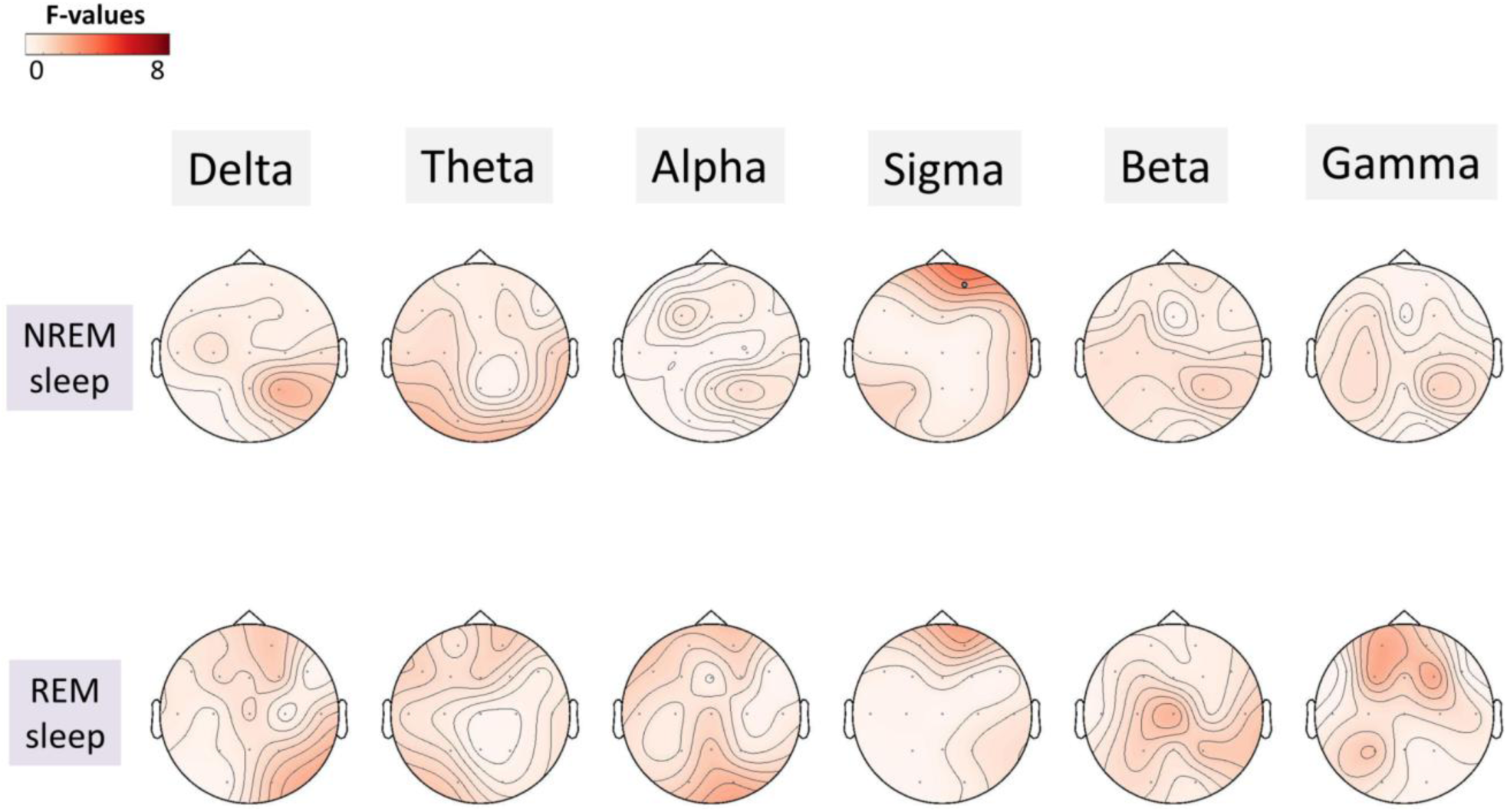
Between-group differences in changes in EEG power following the intervention. Statistical maps (F-values) of the Group × Visit interaction effects on EEG power during NREM and REM sleep. Gray dots (•) indicate channels with p-value < 0.05 at the channel level that did not reach significance after applying the cluster-based permutation approach. Abbreviations: NREM (Non-Rapid Eye Movement), REM (Rapid Eye Movement).

## Discussion

This study examined the effects of an 18-month meditation-based intervention on both subjective and objective sleep parameters in 120 cognitively unimpaired older adults. This intervention was compared with non-native language training (active control) and a no-intervention group (passive control). Questionnaires, actigraphy, and gold-standard polysomnography provided complementary measures of subjective sleep quality, rest–activity rhythms, sleep architecture and microstructure. Compared to the passive control group, meditation training was associated with subtle improvements in subjective measures related to the ease of morning awakening. No effects were detected for any other subjective or objective sleep measures in this cohort.

Our analyses showed that meditation training was associated with greater ease of morning awakening, as assessed by the LSEQ, when compared with the no-intervention group. This beneficial effect on subjective sleep assessment aligns with previous studies ^13–15^. However, no significant differences were observed between the meditation and active control groups. Moreover, we did not replicate the improvements on overall sleep quality, as assessed by the PSQI, reported previously ^13,14^, nor the reduction of insomnia symptoms assessed using the ISI ^15^. A key difference may lie in the characteristics of the study population. Most studies reporting beneficial effects of meditation on subjective sleep quality included older adults with sleep complaints ^13–15^, or more specifically participants with moderate insomnia ^34,35^. Given the strong links between insomnia and psychoaffective factors, insomnia may be a particularly relevant target for mediation-based interventions. In contrast, sleep disorders driven primarily by underlying pathophysiological processes, such as obstructive sleep apnea or REM sleep behavior disorder, may be less likely to respond to meditation alone.

Consistent with our findings, a previous study conducted in older adults not selected based on sleep difficulties reported no significant improvement in subjective sleep quality following meditation training ^16^. In our sample only 37% of the participants had a PSQI score > 5, the commonly used threshold indicative of poor sleep and the presence sleep disturbances. The limited sleep complaints and minimal insomnia symptoms reported by most participants may therefore account for the very modest effects observed. Overall, the effects of meditation on subjective sleep quality may therefore be more readily detectable in individuals with clinically meaningful sleep complaints, particularly insomnia symptoms.

Regarding objective sleep parameters, our analyses revealed no significant effects of meditation training on rest-activity rhythm, sleep architecture or microstructure. To our knowledge, only one previous RCT has examined the effects of a meditation intervention on objective sleep measures. That study showed no benefit of meditation on sleep architecture compared to an active control group combining sleep hygiene education and exercise program ^15,17^. However, because the study included only an active control group receiving a sleep-focused intervention, any potential differences between groups may have been attenuated. In the absence of a passive control group, we cannot exclude the possibility that both interventions helped prevent or attenuate age-related changes in sleep, thereby obscuring any meditation-specific effects. The present study addressed this limitation by including both an active control group (receiving a foreign language learning intervention unrelated to sleep) and a passive control group (with no intervention). Despite this rigorous design, we did not observe significant effects of meditation on objective sleep parameters. Although a statistically significant increase in spindle density was observed, this result did not survive correction for multiple comparisons.

Overall, our findings suggest that meditation may have only subtle effects on subjective sleep quality in older adults with minimal sleep complaints. Interestingly, cognitive behavioral therapy for insomnia (CBT-I), another non-pharmacological approach targeting the behavioral and psychological factors that contribute to insomnia, has also been shown to primarily influence subjective rather than objective sleep parameters ^36^. CBT-I techniques have been combined with mindfulness exercises in Mindfulness-Based Therapy for Insomnia ^37,38^. However, the benefits of MBTI relative to other mindfulness-based interventions remain unclear, as does the identification of individuals most likely to benefit from these approaches ^38^. Of note, meditation has been proposed to enhance sleep quality through both direct and indirect pathways, notably by reducing psychoaffective symptoms ^8^. However, most studies investigating the effects of meditation on sleep have not accounted for psychoaffective factors, making it difficult to determine the specific pathways through which meditation exerts its effects. In the present study, we examined the effects of meditation on sleep quality in participants without clinically significant psychoaffective symptoms, while controlling for individual differences in trait anxiety and depressive symptoms. All results remained unchanged when these factors were not included as covariates in the analyses, suggesting that psychoaffective factors did not account for the observed benefits of meditation on ease of morning awakening. Further research is needed to assess the effects of meditation training in more diverse populations, including participants with clinically significant psychoaffective symptoms.

As highlighted and discussed in previous reports from the Age-Well RCT ^20,39^ , the study population was not fully representative of the general ageing population, as participants were highly educated and cognitively unimpaired, which may have reduced the likelihood of detecting significant meditation effects. The absence of significant effects of meditation on sleep quality may also be attributable to the modalities of practice during the intervention or the timing at which this intervention was initiated. Indeed, the intervention did not specifically target sleep, and participants were instructed to practice at least 20 minutes with no specific instructions regarding the timing of practice. More intensive daily training, or an intervention specifically designed to apply mindfulness to sleep, for instance through evening practice (before going to bed or falling asleep), may yield greater benefits. In addition, initiating meditation practice earlier in life may also be relevant. Some studies suggest that long-term meditation practice in expert meditators is associated with a greater proportion or duration of deep sleep and REM sleep ^40–42^, as well as reduced nocturnal wakefulness and shorter sleep latency ^40^ in middle-aged and older adults. However, these effects have not been consistently replicated across studies ^43,44^. The potential benefits of meditation on sleep may depend on multiple factors, including cumulative practice duration, session frequency, timing of practice during the day, and age at initiation. Future research should examine these variables to better characterize the impact of meditation training on sleep.

Several strengths of the present study should be highlighted. First, the inclusion of two control groups (one active and one passive) enabled a more rigorous assessment of the specific effects of meditation training on sleep. Second, the intervention period was particularly long (18 months) compared to previous studies, (typically 2 to 12 weeks, ^45^), increasing the likelihood of capturing potential changes in sleep. Another notable strength was the assessment of multiple subjective and objective sleep parameters, allowing for a more comprehensive evaluation of the intervention’s effects. Finally, accounting for individual differences in trait anxiety and depressive symptoms represented another strength of the study, as these are important factors to consider when evaluating the effects of meditation interventions.

However, several limitations should also be acknowledged. First, although the intervention included weekly group sessions and daily home practice, the amount or timing of practice may not have been sufficient to induce measurable effects on sleep quality. Future studies should examine whether practice scheduled close to bedtime yields greater benefits. In addition, our sample comprised healthy, highly educated older adults and was therefore not fully representative of the general population. Participants also reported few sleep complaints, insomnia symptoms, or psychoaffective symptoms, limiting the potential to detect intervention effects. Studies in more diverse and clinically vulnerable populations are therefore warranted.

## Conclusion

This 18-month meditation intervention in older adults with minimal sleep complaint revealed only improvements in ease of morning awakening. Any benefits of meditation on sleep in this population appears thus subtle and limited to self-reported outcomes, with no detectable effects on objective sleep measures. These findings help clarify the potential effect of meditation on sleep in aging. Future research should include more diverse populations (particularly individuals with clinically significant sleep disturbances) and apply similar approaches combining subjective and objective sleep assessments, appropriate active and passive control groups, and interventions of sufficient duration.

## Author contributions

PC and AH contributed equally to data collection, data curation, formal analysis, and the writing of the original manuscript. SR contributed to data collection and data curation. FB, OH, and MD contributed to data collection. GC contributed to conceptualization, funding acquisition, and manuscript editing. AL contributed to conceptualization and funding acquisition. AM provided supervision for the analyses and edited the manuscript. GR contributed to conceptualization, funding acquisition, project administration, supervision of the project and the analyses, and the reviewing and editing of the manuscript. All authors have read and agreed to the published version of the manuscript.

## Clinical Trial Information

<u>Name</u>: Study in Cognitively Intact Seniors Aiming to Assess the Effects of Meditation Training (Age-Well).

<u>URL</u>: https://clinicaltrials.gov/ct2/show/NCT02977819?term=Age-Well&draw=2&rank=1.

<u>Registration</u>: EudraCT: 2016-002441-36; IDRCB: 2016-A01767-44; ClinicalTrials.gov Identifier: NCT02977819.

## Acknowledgments

We thank the participants of the study for their contribution. We are grateful to the Cyceron staff for their help with recruitment, and to Aurélia Cognet, Valérie Lefranc and Géraldine Poisnel for their administrative support.

## Data Availability

Data are made available on request following a formal data sharing agreement and approval by the consortium and executive committee (https://silversantestudy.eu/2020/09/25/data-sharing). The Material can be mobilized, under the conditions and modalities defined in the Medit-Ageing Charter, by any research team belonging to an Academic for carrying out a scientific research project relating to the scientific theme of mental health and well-being in older people. The Material may also be mobilized by non-academic third parties, under conditions, in particular financial, which will be established by separate agreement between Inserm and by the said third party. Data sharing policies described in the Medit-Ageing Charter are in compliance with our ethics approval and guidelines from our funding body.

