## Supplementary files for "Effects of an 18-month meditation training on subjective and objective sleep quality in older adults: a secondary analysis of the Age-Well randomized controlled trial"

**Table S1.** Baseline characteristics in the 3 groups of participants.

**Table S2.** Sleep questionnaires scores at baseline and post-intervention according to intervention group and visit.

**Table S3**. Rest-activity rhythm parameters at baseline and post-intervention according to intervention group and visit.

**Table S4**. Sleep architecture parameters at baseline and post-intervention according to intervention group and visit.

**Figure S1.** Between-group changes in spindle density over centroparietal derivations, after 18 months of intervention in each group.

**Table S1. Baseline characteristics in the 3 groups of participants.**

| Variable | Meditation training (n = 40) | Non-native language training (n = 41) | No intervention (n = 39) |
|---|---|---|---|
| Age, year | 69.59 (3.79) [65.28:78.41] | 69.94 (4.47) [65.00:83.86] | 68.01 (2.42) [65.01:75.27] |
| Women, number (%) | 28 (70) | 24 (59) | 24 (62) |
| Education, year | 13.25 (3.14) [7:22] | 12.37 (3.09) [7:17] | 14.13 (2.67) [7:20] |
| MMSE | 28.95 (1.20) [26:30] | 29.07 (0.98) [27:30] | 29.21 (0.92) [27:30] |
| STAI-B | 36.35 (7.47) [20:50] | 34.68 (7.39) [20:54] | 33.13 (6.74) [23:50] |
| GDS | 1.55 (2.07) [0:11] | 1.12 (1.52) [0:6] | 1.21 (1.76) [0:7] |
| Apnea-Hypopnea Index, number of events/h | 25.33 (16.21) [6.40:62.10] | 27.03 (16.26) [2.40:75.50] | 21.79 (11.90) [0.70:54.00] |

Continuous data are presented as mean (SD) [range]. *Abbreviations: GDS (Geriatric Depression Scale), MMSE (Mini-Mental State Examination), SD (standard deviation), STAI-B (State-Trait Anxiety Inventory B).*

**Table S2. Sleep questionnaires scores at baseline and post-intervention according to intervention group and visit.**

| | Meditation training | | Non-native language training | | No intervention | |
|---|---|---|---|---|---|---|
| **Questionnaire** | Baseline (n=40) | Post-intervention (n=40) | Baseline (n=41) | Post-intervention (n=41) | Baseline (n=39) | Post-intervention (n=38) |
| PSQI | 5.12 (3.50) | 5.42 (3.51) | 4.88 (3.25) | 5.32 (3.42) | 4.62 (2.91) | 4.50 (2.67) |
| PSQI > 5 (n) % | 17 (42.5%) | 19 (47.5%) | 14 (34.2%) | 16 (39.0%) | 13 (33.3%) | 9 (28.7%) |
| ISI | 7.05 (6.00) | 7.04 (6.00) | 6.49 (5.35) | 6.77 (5.54) | 6.54 (4.34) | 6.12 (4.60) |
| ESS | 4.58 (2.58) | 4.67 (2.80) | 5.49 (3.61) | 5.46 (2.66) | 5.28 (3.84) | 5.55 (3.89) |
| LSEQ getting to sleep | NA | 0.32 (1.26) | NA | 0.33 (0.93) | NA | 0.01 (0.72) |
| LSEQ quality of sleep | NA | 0.29 (1.72) | NA | 0.21 (1.40) | NA | 0.02 (1.13) |
| LSEQ awakening from sleep | NA | 0.40 (1.30) | NA | 0.09 (1.12) | NA | -0.08 (0.78) |
| LSEQ behavior following wakefulness | NA | 1.43 (1.63) | NA | 1.16 (1.62) | NA | 0.90 (1.38) |

*Scores on sleep and wakefulness questionnaires are presented as mean (SD). Abbreviations: ESS (Epworth Sleepiness Scale), ISI (Insomnia Severity Index), LSEQ (Leeds Sleep Evaluation Questionnaire), NA (not applicable), PSQI (Pittsburgh Sleep Quality Index), SD (standard deviation).*

**Table S3. Rest-activity rhythm parameters at baseline and post-intervention according to intervention group and visit.**

| | Meditation training | | Non-native language training | | No intervention | |
|---|---|---|---|---|---|---|
| **Rest-activity rhythm** | Baseline (n=38) | Post-intervention (n=37) | Baseline (n=40) | Post-intervention (n=37) | Baseline (n=37) | Post-intervention (n=37) |
| Interdaily stability | 0.92 (0.04) | 0.91 (0.04) | 0.92 (0.04) | 0.92 (0.04) | 0.92 (0.05) | 0.92 (0.04) |
| Intradaily variability | 0.62 (0.11) | 0.62 (0.07) | 0.64 (0.11) | 0.63 (0.09) | 0.67 (0.47) | 0.57 (0.11) |
| Relative amplitude | 0.79 (0.15) | 0.80 (0.14) | 0.86 (0.25) | 0.78 (0.22) | 0.83 (0.21) | 0.77 (0.22) |

*Rest-activity rhythm parameters are presented as mean (SD). Abbreviation: SD (standard deviation).*

**Table S4. Sleep architecture parameters at baseline and post-intervention according to intervention group and visit.**

| | Meditation training | | Non-native language training | | No intervention | |
|---|---|---|---|---|---|---|
| **Sleep architecture** | Baseline (n=40) | Post-intervention (n=40) | Baseline (n=41) | Post-intervention (n=41) | Baseline (n=39) | Post-intervention (n=38) |
| TST (min) | 361.11 (63.20) | 331.62 (54.93) | 352.38 (70.22) | 357.36 (57.31) | 366.55 (61.72) | 373.22 (59.94) |
| Sleep efficiency (%) | 76.72 (9.71) | 74.69 (11.78) | 74.82 (10.21) | 78.63 (10.75) | 79.26 (10.18) | 82.24 (9.50) |
| Sleep latency (min) | 19.80 (13.34) | 19.69 (15.70) | 23.93 (14.92) | 17.83 (14.83) | 19.18 (13.52) | 16.49 (27.14) |
| WASO (min) | 91.20 (49.77) | 94.40 (51.44) | 45.81 (13.83) | 81.53 (50.19) | 47.10 (11.79) | 64.51 (38.80) |
| NREM-1 (% TST) | 14.97 (9.10) | 11.40 (7.78) | 13.83 (7.46) | 9.76 (5.51) | 11.79 (6.01) | 8.88 (4.34) |
| NREM-2 (% TST) | 48.11 (8.21) | 52.85 (8.03) | 49.52 (8.57) | 53.52 (8.59) | 47.90 (8.34) | 50.83 (8.93) |
| NREM-3 (% TST) | 18.01 (9.31) | 18.37 (8.58) | 19.62 (9.67) | 19.72 (8.82) | 21.83 (9.51) | 21.79 (7.14) |
| REM (% TST) | 18.89 (4.54) | 17.40 (5.28) | 17.04 (6.51) | 17.00 (5.85) | 18.48 (5.55) | 18.82 (5.00) |

Sleep architecture parameters are presented as mean (SD). *Abbreviations: NREM (Non-rapid eye movement sleep), REM (rapid eye movement sleep), SD (standard deviation), TST (Total Sleep Time), WASO (wake after sleep onset).*

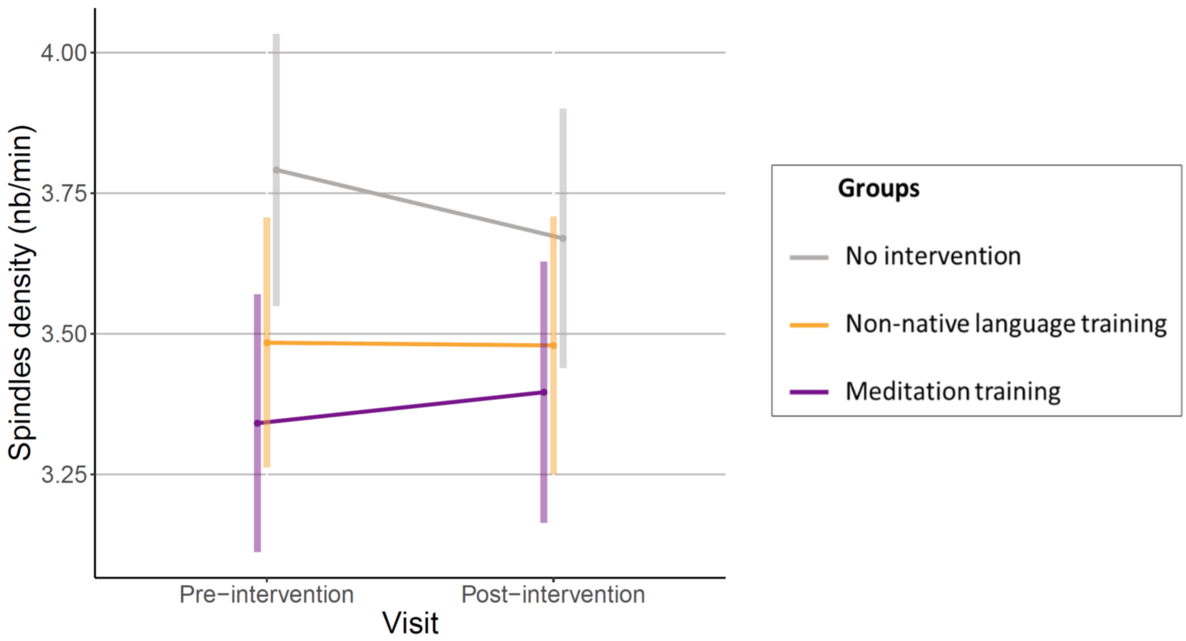


**Figure S1. Between-group changes in spindle density over centroparietal derivations, after 18 months of intervention in each group**.

*Spindle density was averaged across centroparietal channels (C3, Cz, C4, P3, Pz, P4). Error bars indicate 95% confidence intervals around the estimated marginal means.*
